# Conjunctival polymerase chain reaction-tests of 2019 novel coronavirus in patients in Shenyang, China

**DOI:** 10.1101/2020.02.23.20024935

**Authors:** Li Xu, Xinyue Zhang, Wei Song, Baijun Sun, Jinping Mu, Bing Wang, Zhiqian Wang, Yehong Cao, Xue Dong

## Abstract

**Purpose:** The 2019 novel coronavirus(COVID-19) mainly transmitted by person-to-person through inhalation of respiratory droplets. We report the laboratory results of conjunctival PCR-tests and some clinical features of these patients in shenyang China.

**Design:** This is a cross-sectional non-randomized study

**Subjects:** The study include 14 confirmly diagnosed cases, 16 suspected cases and some medical observed patients.

**Methods:** All patients with diagnosed and suspected COVID-19 were admitted to a designated hospital in Shenyang, China. We collected conjunctival samples of these patients to do the laboratory tests by real time RT-PCR. Medical observed patients were enrolled if they had clinical symptoms. Then we analysed the PCR results and clinical data from eletronic medical records in order to find some relationships.

**Main Outcome Measures:** Clinical condition and PCR results. of conjunctival swabs compared with other specimens

**Results:** One of the identified case coverted from suspected case without typical clinical symptoms. Twenty-two medical observed cases were removed because none of them converted to identified cases. One of the suspected converted to identified case recently. The included cases in our study are imported cases with less underlying diseases and the severity of their infection was relatively moderate. All the conjunctival results of PCR-test were negative. Two cases had typical clinical symptoms but were finally confirmed by repeated pharynxswabtests.

**Conclusion:** Conjunctiva may be a transmission way of COVID-19. And ocular conjunctival swabs in combination with PCR test could be a non-invasive, convenient and feasible diagnostic method for identifying the infection of COVID-19. Emphasis on the false-negative results is vital.

## Introduction

Recently, a new strain of coronavirus, designated as COVID-19 by WHO, firstly emerged in Wuhan, Hubei, China, and spreaded to multiple cities rapidly. Coronaviruses are enveloped non-segmented positive-sense RNA viruses belonging to the family Coronaviridae and the order Nidovirales, which are widely detected in humans and other mammals, and commonly denoted in etiologies of respiratory tract infections in humans [1-3].With similarity to SARS and MERS, they are all dangerous zoonotic coronaviruses which may have the potential to cause a threatening panepide micamong human beings [4-5].

Based on clinical experience, COVID-19 is mainly disseminated by person-to-person through inhalation of respiratory droplets. However, some researchers reported that the viruses could also transmitted by direct contact[6] and digestive tract [7], and conjunctiva may also be possible transmission route without validation [8]. As previously reported, two clinical doctors were also infected with COVID-19 who wore N95 masks during his work, but they did not wear gogglesat that time leaving their ocular surface directly to the room air.

In this study, we make an effort to testify whether conjunctiva is a possible way for the transmission of COVID-19 and whether it could be an effective diagnostic approach to detecting the infection of COVID-19.

## Methods

### Data Collection

In this cross-sectional non-randomized study, 14 identified cases, 16 suspected cases and 22 medical observed cases that infected with from January, 2020 to February, 2020 were included. The study was approved by local ethics committee, sponsored by Ministry of Shenyang science and technology and in accordance with Declaration of Helsinki. At the time of collecting conjunctival samples, every patient was informed of purposes and methods, and informed consent was obtained from all patients. Fundamental information on age, gender, underlying systemic disease and severity of diseases. Ophthalmic information on ocular signs and symptoms, history of ocular surgery and the results of conjunctival PCR-tests were recorded. Detailed research methods were listed as follows.

### Nucleic acid extraction and real time RT-PCR

After obtaining conjunctival samples by sterile swabs, the samples were placed in EP tubes filled with virus sampling fluid. Then the samples were transported immediately to the −80°C refrigerator in order to preserve and isolate COVID-19 subsequently.

### Nucleic acid extraction and PCR analysis

Specimens were extracted using a viral nucleic acid detection kit(QIAGEN). Briefly, 200μL sample was used to prepare reaction mixture. After 30 minutes’standing, the reaction mixture was centrifuged for 10 seconds. The reaction system consisted of 13 μL Cov test buffer and 5μL reaction fluid. After extracting the viral DNA, then the samples were used for COVID-19 PCR analysis. The assay was performed in the laboratory with a real-time Light Cycler (ABI7500).Reactions were set up performed according to the manufacturer’s instructions. Real-time PCR was performed using the Light Cycler and targeted agents(made by Jienuo Biotechonology Limited Company in Shanghai) to amplify the COVID-19 at 42 °C for 10s, followed by 95°C for 10s, and finally 40 cycles for 10s at 95 °C and for 45s at 60 °C.

The results mainly depended on the CT values of two targets, the ORF(Open reading frame) and core-shell protein gene.

## Results

We retrospectively analyze 30 patients who were diagnosed as infection of COVID-19, including 14 identified cases and 16 suspected cases. One suspected case has coverted to identified case recently. What’s more,22 medical observed cases who had had intimate contact with identified or suspected cases were included. However, because none of medical observed cases have converted to identified cases and related detection of samples were negative, so they are removed from this study. As displayed in the table 1, the mean age of the identified group is 48±13.4 years old and that of the suspected group is 40±16. 2 years old. The numbers of male and female are nearly equal. As for systematic diseases,3 of them have diabetic mellitus, 4 of them have hypertension and 1 of them has hepatitis B. None of them have the history of general or ocular surgery. Just 1 identified case is complicated with macular degeneration and another 1 identified case felt eye itching before the onset.

**Table 1:** Demographics and baseline characteristics of identified and suspected cases infected with COVID-19

|  | All patients(N=30) | Identified(n1=14) | suspected(n2=16) |
| --- | --- | --- | --- |
| Characteristics |  |  |  |
| Age, years | 43.7 ± 14.3 | 48 ± 13.4 | 40 ± 16.2 |
| Sex |  |  |  |
| Men | 14 ( 46.7% ) | 7 ( 50% ) | 7 ( 43.75% ) |
| Women | 16 ( 53.3% ) | 7 ( 50% ) | 9 ( 56.25% ) |
| Earliest Contact Time |  | 2020.01.04 | 2020.01.13 |
| Last Contact Time |  | 2020.01.31 | 2020.01.31 |
| Onset Time,numbers |  |  |  |
| January | 16 ( 55.2% ) | 10 | 6 |
| February | 13 ( 44.8% ) | 4 | 9 |
| Identifying Time,numbers |  |  |  |
| January | 4 ( 18.2% ) | 4 | 0 |
| February | 18 ( 81.8% ) | 10 | 8 |
| Underlying Systematic Disease |  |  |  |
| Diabetes | 3 ( 37.5% ) | 3 | 0 |
| Hypertension | 4 ( 50% ) | 4 | 0 |
| Hepatitis | 1 ( 12.5% ) | 1 | 0 |
| Ocular or General Surgery History | None | None | None |
| Ocular Symptoms |  |  |  |
| eye itching | 1 | 1 | 0 |
| Ocular Comorbidity |  |  |  |
| macular degeneration | 1 | 1 | 0 |
| Severity of Disease | (8 cases were removed from suspected group without abnormalities) |  |  |
| mild | 6(27.3%) | 2 | 4 |
| moderate | 3(13.6%) | 2 | 1 |
| ordinary | 4(18.2%) | 3 | 1 |
| severe | 1(4.5%) | 1 | 0 |
| common pneumonia | 6(27.3%) | 5 | 1 |
| severe pneumonia | 2(9.1%) | 1 | 1 |
| Total | 22(100%) | 14(63.6%) | 8(36.4%) |

With regard to the severity of disease, common pneumoniae accounts for most of identified cases and mild cases are the most common type of the suspected cases. In the identified group, there is just one severe pneumonia case, whereas another one converted to severe pneumonia from a suspected case.

The test results and related time show as below in table 2.

**Table 2:**
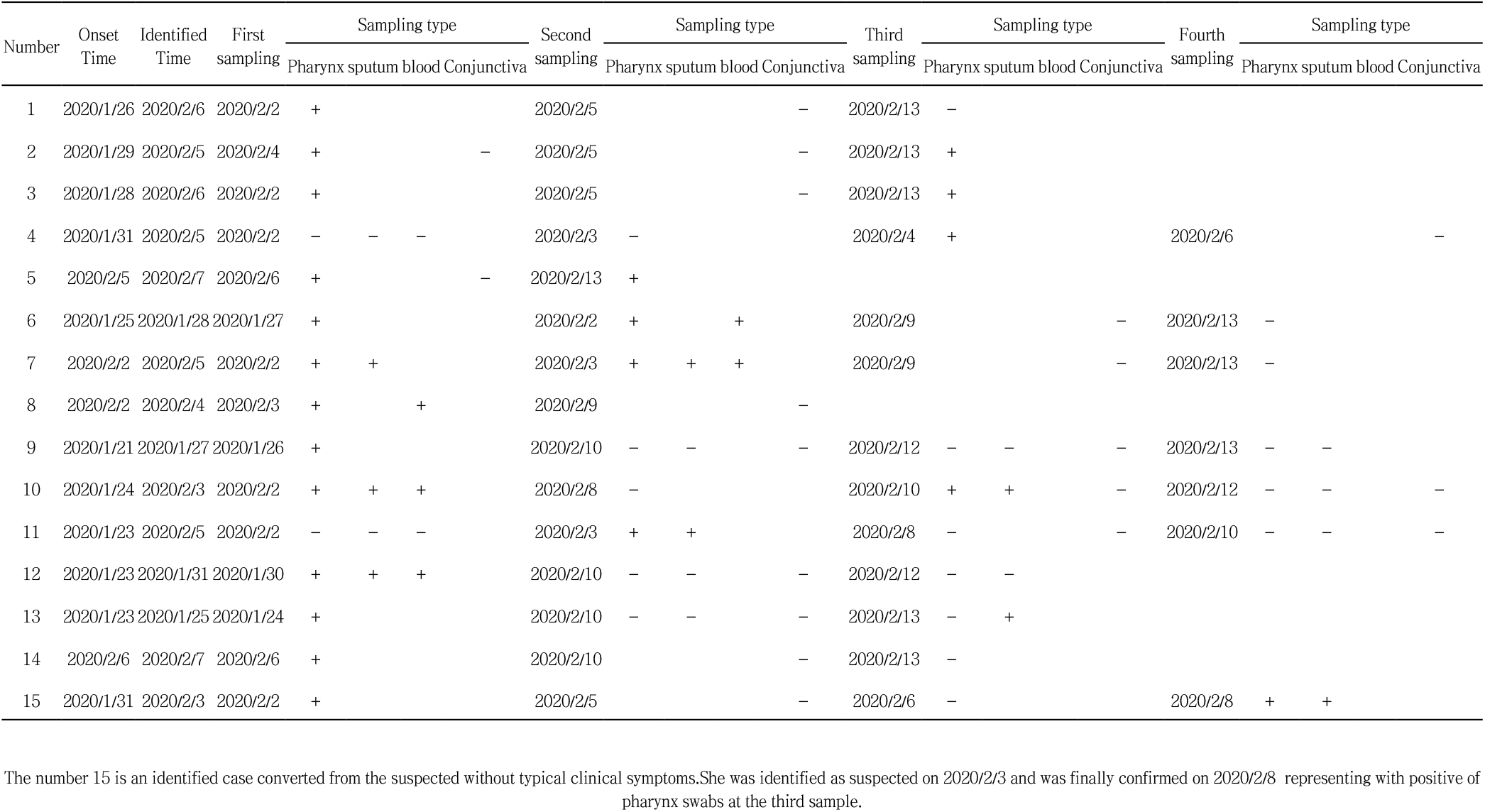
The detection results of identified patients with infection of COVID-19

## Disscussion

Generally speaking, deducted from the fundamental condition of our cases, we found that the severity of disease in our patients is relatively moderate. In addition, the affected age group is younger without so many underlying diseases. There are only 2 cases manifested as severe pneumonia, and one of them coverted from suspected case. By investigating the possible reasons, we speculated this consequence may cause by the delayed confirmation of infection. However, the condition seems to contradict the epidemiological study done by a cluster of Chinese researchers, in which they found that COVID-19 infection could cause severe respiratory illness similar to severe acute respiratory syndrome and was associated with ICU admission and high mortality[9]. It may be due to the fact that the included are all imported cases who recently have gone to Wuhan or contacted with the identified cases, compared with those who were invaded directly by viruses.

PCR test has been widely used to confirm the viral infection targeted to the virus-specific nucleic acid sequences, but in our study, we found that after isolation for presumed infection of COVID-19, the 2 cases were finally diagnosed by repeated pharynx swabstests. We postulated that may beit caused by false sampling position, which means that the virusesare likely to exist in lower respiratory tract other than the upper. So the false-negative rate of PCR may mislead the clinical doctors and even miss the optimal opportunity for treatment. Some Chinese clinicians also found similar phenomenon and reported 5 cases. By evaluating radiographic characteristics of 5 patients with confirmed 2019-nCoV infection and initial negative or weakly positive

RT-PCR, They found that the patients presented characteristic radiographic features of COVID-19 pneumonia from the first scan and then were confirmed by positive repeat swabs test during the isolated observation or treatment. They ascribed the possible reasons to laboratory error or insufficient viral material in the specimen[10].

In our cases, all of the conjunctival results of PCR were negative no matter in identified or suspected cases. We analyze and summarize the reasons as follows: (1) The shedding loads of the COVID-19 was below the sensitivity of the test or some individuals were actually not shedding viral DNA at that time. This opinion correlates with Sarrah E. Burr’s study, where she obtained 28 conjunctival swabs during the outbreak of acute haemorrhagic conjunctivitis in the Gambia, West Africa, caused by the epidemic of coxsackievirus, and 25% of the PCR-test showed negative results. [11] (2) In our study, the conjunctival sampling time of the cases was after identification. The administration of drugs, such as corticosteroid and antiviral drugs, and the mutation of viruses could all affect the results. (3)The sensitivity of the viral nucleic acid kits is low. (4) The conjunctiva lacks of related receptors. A pivotal factor for efficient person-to-person transmission is the ability of the virus to attach to human cells. Because coronaviruses use a spike protein for attachment to host cells[12]. As previously reported, both COVID-19 and SARS-nCov use the same receptor called ACE2, which has been verified to locate mainly on lung alveolar epithelial cells and enterocytes of the small intestine[13]. However, whether ACE2 receptors exist on the conjunctival surface and the level of conjunctival ACE2 expression still need further investigation. Given that ACE2 expression is extremely rare on the ocular surface, the viruses could not attach to the conjunctiva and they may transfer to any organs in our bodies through lacrimal ductule. In our study, a 29-year-old female without any systematic disease converted to identified case from suspected case. She was a moderate suspected case, whose result of pharynx swab was positive at first, and after treatment, the result changed negative. However, the result represented as positive after 3 days. So we could speculate present therapies may only resist the viruses temporarily, and the COVID-19 may seek any opportunity to represent when the human immunity weakens. What’s worse, they may transfer to anywhere to find suitable hosts. In addition, we found that the sampling time contributed to obtain a reliable result, pharynx and conjunctiva swab should be collected meantime. In our study, the two swabs were obtained meantime in 3 identified cases, but the results were inconsistent. The coexistence of positive pharynx specimen and negative result of conjunctiva specimen reminded that the viruses may be more likely to attach to respiratory tract other than conjunctiva.

Restricted by reality, there are some limitations our study. Firstly, no positive result was obtained from identified and suspected cases. Secondly, we lack of swabs collected from individuals with normal eyes during the sampling process.

Undoubtedly, the regular samples for PCR tests, such as pharynx and sputum swabs, have its advantage in some facets, but considering their errors and the viral transmission characteristics, conjunctival swabs for PCR may become a convenient, noninvasive and simple diagnostic methods. And whether conjunctiva is a possible transmission route of COVID-19 still needs further research. Sampling time is also a key factor as well. A more uniformed scheme should be established to guide the clinical practitioners make correct judgements avoiding the cross infection.

## Data Availability

the data are all in the manuscript

## Abbreviation

RT-PCR: reverse transcription polymerase chain reaction
COVID-19: coronavirus disease 2019

